# Allostatic Load in Endometrial Cancer Disparities

**DOI:** 10.64898/2026.06.06.26355062

**Authors:** Ganga S. Bey, Mikayla Borthwick Bowen, Sirui Wu, Matthew Boykin, Laurence Bernard, Qian Zhang, Brenda Melendez, Joseph Celestino, John A. Batsis, Charlotte Sun, Feng-Chang Lin, Melinda S. Yates

## Abstract

**Background:** Endometrial cancer incidence and mortality are increasing, particularly among Black women and for aggressive subtypes. Allostatic load (AL), a composite measure of physiologic dysregulation across metabolic, cardiovascular, and immune systems, varies by racial category and tumor subtype in other cancers. Endometrial cancer is strongly associated with obesity, and it is unknown whether AL scores maintain sufficient heterogeneity to evaluate differences across subgroups or with clinical outcomes.

**Objective:** To describe the performance of AL scoring in endometrial cancer patients and examine associations with tumor characteristics (grade/histology) and survival outcomes.

**Methods:** We evaluated AL among 398 participants newly diagnosed with endometrial cancer. AL score was calculated by assigning 1 point for each “high-risk” value (by clinical reference range or distribution-based) for 15 biologic variables for vital signs, anthropometrics, blood-based biomarkers, and medical comorbidities.

**Results:** Distribution-based thresholds for variables were used to preserve heterogeneity in this obesity-dominant context. Overall, 68.7% of Black women had high AL compared to White (56.7%), Hispanic (56.7%), and other race (32.3%) women. Decision tree analyses revealed grade-dependent associations between AL and survival. For women with low-grade tumors, higher AL was associated with poorer overall survival. For high-grade tumors, intermediate AL (≥4, <8) were associated with shortest overall survival. Black women with low-grade disease experienced shorter progression-free survival regardless of AL.

**Conclusions:** AL scoring maintains heterogeneity despite high obesity prevalence in endometrial cancer. Varying relationships between AL and survival by tumor grade and ethnoracial group suggest cumulative physiologic burden and social/structural factors may jointly shape endometrial cancer disparities.

## 1. Introduction

Endometrial cancer is one of few cancers in the United States marked by increasing incidence and mortality [1]. While obesity is one of the strongest predictors, increasing rates of obesity do not completely explain the sharp increase in incidence of endometrial cancer [2]. Endometrial cancer rates increased approximately 1% per year from 2003 to 2015, with the most rapid increases seen in Hispanic, Asian, and non-Hispanic Black (“Black”) women [2]. Additionally, rate increases were most pronounced for more aggressive, non-endometrioid subtypes, including serous, clear cell, and carcinosarcoma, which comprise 18.3% of all endometrial cancers [2, 3]. These aggressive subtypes are twice as frequent in Black women, who experience lower overall five-year survival across all stages and subtypes of endometrial cancers compared to non-Hispanic White (“White”) women [2].

In explaining Black women’s poorer health outcomes, researchers have proposed the “weathering” hypothesis, which posits that early health deterioration within this group is a consequence of the unique cumulative impact of repeated experiences with social and economic adversity and marginalization stemming from interlocking systems of racial and gender inequities [4, 5]. On a physiological level, persistent, high-effort coping with acute and chronic stressors is known to have a profound effect on health [6, 7]. A well-established literature base has linked social adversity to these underlying physiological mechanisms hypothesized to yield racial disparities in health. Cortisol levels, blood pressure, cytokine production, waist-to-hip ratio, and glycated hemoglobin levels have been associated with socioeconomic status, occupation, birth outcome, and environmental risk [8].

Consistent with the weathering hypothesis, some of the earliest work linking social environmental stressors to cumulative physiological burden leveraged the concept of allostatic load, or wear and tear on the body’s systems owing to repeated adaptation to stress [9]. Allostatic load has been operationalized as an index that captures cumulative physiologic effects (including the body’s response to chronic physiologic stress) across major regulatory systems, resulting from dysregulation of primary mediators in the hypothalamic pituitary adrenal (HPA) axis [10, 11]. Across studies, Black women have consistently been demonstrated to have a higher age-adjusted allostatic load than Black men and White women [8, 12, 13]. Substantial evidence has linked this dysregulation to several downstream health effects, including cardiovascular and metabolic conditions that lead to increased risk for a wide variety of chronic diseases, including cancers [9, 14, 15].

Racially patterned disparities in allostatic load may play a role in racial disparities in cancer. Mechanistically, allostatic load reflects sustained activation of neuroendocrine, metabolic, inflammatory, and immune pathways, systems that are central to carcinogenesis and tumor progression [14]. Under conditions of chronic psychosocial stress (e.g., persistent exposure to inequity-driven cultural, economic, or psychological stressors), prolonged disruptions in homeostasis may promote a pro-tumorigenic microenvironment characterized by insulin resistance, systemic inflammation, immune dysfunction, and altered steroid hormone signaling. Similar pathophysiologic mechanisms plausibly underlie a relationship between elevated allostatic load and adverse cancer outcomes. These biologic perturbations may contribute to more aggressive tumor behavior and poorer survival outcomes.

A growing body of literature suggests associations between elevated allostatic load and adverse cancer outcomes. Triple-negative breast cancer (TNBC) is an aggressive subtype of breast cancer with poor prognosis [16]. Black women are twice as likely to develop TNBC compared with White and Hispanic women [17]. Studies examining racial disparities in breast cancer have shown that allostatic load is correlated with poor prognostic characteristics, particularly in Black women [18–20]. An analysis of National Health and Nutrition Examination Survey (NHANES) showed that allostatic load is significantly associated with increased breast cancer risk among Black women but not White women [21]. While the epidemiologic profile of aggressive endometrial cancer subtypes somewhat mirrors that of TNBC, the relationship of allostatic load with endometrial cancer has not yet been empirically evaluated.

Obesity is disproportionately higher among Black women compared with White women [22] and is estimated to account for approximately 57% of endometrial cancers in the United States [23, 24]. However, studies of disparate cancer risk and survival outcomes related to obesity have often lacked evaluation of different impacts across low-grade endometrioid, high-grade endometrioid, and non-endometrioid subtypes along with representation across racial/ethnic groups and other social factors. In addition, the degree of obesity and metabolic dysfunction are likely contributors to these risks but are often collapsed into binary (obese versus non-obese) categories based on body mass index (BMI).

BMI is an imperfect anthropometric measure that fails to capture obesity-associated metabolic, inflammatory, and neuroendocrine heterogeneity, processes that overlap substantially with biologic domains encompassed by allostatic load. Studies by our team and others have indicated that a convergence of obesity with additional physiologic stressors, such as insulin resistance, inherited cancer susceptibility, immune response, and other factors may contribute to obesity-driven reprogramming in both premalignant endometrium and in endometrial tumors [23, 25–31]. Collectively, these findings suggest that allostatic load may represent a biologically integrative framework linking chronic stress exposure, metabolic dysfunction, and endometrial carcinogenesis, particularly in populations experiencing disproportionate cumulative stress burden.

Evaluating allostatic load as a biologically integrative framework to disentangle the complex, interacting mechanisms contributing to the rising incidence and mortality of endometrial cancers could be an essential step toward identifying effective individual- and population-level intervention strategies. Because obesity alone does not adequately account for the increasing burden of endometrial cancer, particularly non-endometrioid and other high-risk subtypes, our first goal was to evaluate allostatic load score performance for women with endometrial cancer. We then sought to evaluate whether elevated allostatic load is associated with adverse tumor clinicopathologic features and poorer survival outcomes in women diagnosed with endometrial cancer, with specific attention focused on the impacts across tumor subtypes and groups defined by race and ethnicity. Specifically, we hypothesized that: 1) allostatic load can meaningfully describe heterogeneity in multisystem physiological burden even within the context of an obesity-associated malignancy, irrespective of race/ethnicity; and 2) high allostatic load is associated with poorer clinical outcomes (progression-free survival and overall survival) in endometrial cancer.

## 2. Methods

### 2.1 Study design and participants

This study utilized clinical data and blood-based assay data from patients with previously untreated, newly diagnosed, and histologically confirmed stage I to III endometrial cancer at MD Anderson Cancer Center during 2016 to 2022. All participants provided consent to provide biospecimens to the Gynecologic Oncology Tumor Bank. This research protocol was approved by the institutional review boards at MD Anderson Cancer Center and University of North Carolina (for data analysis). Patients with other concurrent cancers, with recent or concurrent chemotherapy and/or radiation therapy (within the previous 12 months), and patients with stage IV endometrial cancer were excluded. Clinical data were abstracted from the electronic health record and blood-based biomarker assays were conducted using archived banked specimens from the Gynecologic Oncology Tumor Bank at MD Anderson. Only blood samples collected prior to initiating treatment were evaluated. Inclusion of participants consented to the Gynecologic Oncology Tumor Bank during 2016 – 2022 were planned to target a total sample size of 400 cases. To reach the total analytic sample with oversampling of aggressive disease, a consecutive series of participants with aggressive tumor subtypes (high-grade/non-endometrioid) in this timeframe were included (reaching n=168). In parallel, a consecutive series of cases with less aggressive subtypes (low-grade endometrioid) were included to reach the total sample size.

### 2.2 Data collection

Patient information was abstracted from each patient’s medical records at MD Anderson Cancer Center, including race, ethnicity, age at diagnosis, past medical history (e.g. diabetes mellitus, hypertension, current use of medications for hypertension or diabetes), social history (e.g. smoking status, employment status, etc.), gynecologic and obstetrical history (e.g. parity, use of hormonal menopausal therapy), family history of cancer, physiological data at the first consultation prior to treatment (e.g. blood pressure, resting heart rate), FIGO stage, measured body mass index (BMI) at diagnosis, and treatment type for endometrial carcinomas including treatment or surgery and oncologic outcomes (progression-free survival and overall survival). Waist circumference was determined based upon diagnostic/pre-operative CT scans using previously described methods [32, 33]. Briefly, the cross-sectional image at the midpoint of the L3 spinal region was imported to ImageJ for each patient, where the threshold was adjusted to a lower limit of “250” and upper limit of “1000” to aid in automatic tracing of the body perimeter. The wand (tracing) tool was used to select the highlighted image and measure the perimeter (waist circumference).

### 2.3 Study variables

#### Exposure: Allostatic Load

As there is no consensus on a single operationalization of allostatic load, scores were determined using 15 previously reported variables representing both primary and secondary mediators [8, 20, 21, 34–36] based on clinical information or blood-based biomarkers. For each biomarker in which patients’ values exceeded threshold values (described in detail in statistical analyses below), one point was assigned. Variables included systolic blood pressure (SBP), diastolic blood pressure (DBP), high-density lipoprotein (HDL), total cholesterol, triglycerides, waist circumference, BMI, resting heart rate (RHR), hemoglobin A1c (HbA1C), albumin, c-reactive protein (CRP), interleukin-6 (IL-6), estimated glomerular filtration rate (eGFR), creatinine, fasting glucose, and ever use of medications to control hypertension, diabetes, or hypercholesterolemia. Clinical reference range threshold values were defined as follows: (1) SBP ≥ 140 mmHg; (2) DBP ≥ 90 mmHg; (3) HDL < 50 mg/dL; (4) total cholesterol > 240 mg/dL or total cholesterol < 240 mg/dL and LDL > 130 mg/dL; (5) triglycerides > 150 mg/dL; (6) waist circumference > 88 cm; (7) BMI > 30; (8) RHR > 100 bpm; (9) HbA1C > 6.5; (10) albumin < 4 g/dL; (11) CRP > 3 mg/L; (12) IL-6 > 1.8 pg/mL; (13) eGFR < 60 mL/min/1.73m^2^; (14) Creatinine > 1.2 mg/dL; (15) fasting glucose ≥ 110 mg/dL. Thresholds based on sample distributions are described below and in the results section. Scores were calculated as a summary of assigned points, with maximum possible score of 15 (range: 0–15).

#### Clinical outcomes

The co-primary clinical outcomes in our analyses were progression-free survival and overall survival. Survival outcome start dates were set as the date of diagnosis (e.g., at the time of endometrial biopsy or surgery). For progression-free survival, the end dates were set as the date of recurrence, disease progression, or death. For overall survival, the end dates were set as the date of death. Those alive were censored at their date of last documented contact in the medical record.

#### Tumor characteristics

Details of endometrial cancer histology and grade were abstracted from the clinical pathology report of the hysterectomy specimen for each participant. Tumor histology included endometrioid, serous, clear cell, carcinosarcoma, undifferentiated, or mixed histology (when more than one histologic subtype was identified on the clinical pathology report). In the absence of molecular subtyping data, grade and histology were used in our analysis for defining tumor subtype for additional statistical analyses. Individual histologic categories (“5-category” analysis below) included endometrioid, serous, clear cell, carcinosarcoma, or mixed histology. Alternatively, tumors were further grouped into endometrioid and non-endometrioid groups (described as “2-category” analysis below) to broadly capture major biological and prognostic differences, where non-endometrioid included all serous, clear cell, carcinosarcoma, undifferentiated, or mixed histologies. Tumors with non-endometrioid histology were considered high-grade according to standard practice. Further sensitivity analyses also used the individual histologic subtypes.

Serum and whole blood assay results were abstracted from the electronic health record for each patient from diagnostic/pre-operative samples, including cholesterol, triglycerides, HbA1C, glucose, insulin, eGFR, creatinine, and albumin. Banked serum specimens were utilized to measure interleukin (IL)-6 and C-reactive protein (CRP) at the Proteomics Core at Dan L. Duncan Cancer Center at Baylor College of Medicine. Samples were thawed on ice and centrifuged at 14,000 rpm for 5 minutes to remove debris. The ELLA instrument and cartridge (ProteinSimple, San Jose, CA) were verified using the verification cartridge before use. Samples on the cartridge were analyzed by Simple Plex Runner and Simple Plex Explorer (ProteinSimple, San Jose, CA) was used to obtain and analyze results.

Biomarker and clinical data were collected and managed using REDCap (Research Electronic Data Capture System) hosted at MD Anderson Cancer Center. REDCap is a secure, web-based software platform to support data capture for research studies [37, 38].

#### Race and Ethnicity

Participants self-identified their ethnicity and race from a pre-established list available during routine procedures (not specific to this study). Participants could select Hispanic or non-Hispanic ethnicity, and Black, White, American Indian/Alaska Native, Asian, Native Hawaiian or Other Pacific Islander, or other race. For this analysis, we grouped participants into four categories defined by ethnicity and race: non-Hispanic Black (“Black”), non-Hispanic White (“White”), Hispanic (including Hispanic Black, Hispanic White or Hispanic other race), and non-Hispanic other race, which included participants who identified as non-Hispanic but not Black or White due to limited sample size (n = 31).

#### Social risk score

The Social Vulnerability Index (SVI) is a measure developed by the Centers for Disease Control to identify risk-levels of communities that may be vulnerable before, during, or after large-scale emergencies such as disease outbreaks, natural disasters, or other public health emergencies. The SVI has been found to correlate with several cancer outcomes including cancer stage at diagnosis, surgical outcomes, and mortality [39–44]. SVI score was determined using patient census tract data abstracted from home address according to previously reported methods [45]. We operationalized SVI as both a continuous score ranging from 0 to 1 and sample-based tertiles, where higher scores indicate greater vulnerability.

### 2.4 Statistical analysis

Descriptive statistics were used to summarize patient demographics, clinical characteristics, tumor clinicopathologic characteristics, and allostatic load scores, stratified by ethnoracial group. Continuous variables were reported as means and standard deviations (SD) and compared using analyses of variance (ANOVA). Categorical variables were reported as frequencies and percentages and compared using Chi-squared tests or Fisher’s exact tests when appropriate. The two survival outcomes, progression-free survival and overall survival, were described using Kaplan-Meier curves and compared between groups using log-rank tests. A 95% confidence interval (CI) for the survival function was constructed using Greenwood’s formula for standard error. For data missingness, we used multiple imputations with predictive mean matching (PMM) [46] on the biomarkers to construct the allostatic load scores. A single dataset was generated by averaging across 50 imputed datasets with high stability across imputations.

To define “high-risk” values (used to calculate +1 point for the score) in constructing the allostatic load measure, we then compared two parallel approaches for biomarker thresholds: (1) established standard clinical reference (“clinical”) thresholds and (2) thresholds designated by distribution (“distribution-based”) within our imputed dataset. The distribution-based thresholds were defined as above the 75^th^ percentile for risk-elevating biomarkers (greater than quartile 3, >Q3) and below the 25^th^ percentile (less than quartile 1, <Q1) for HDL, albumin, and eGFR. The allostatic load score was computed as the sum of the dichotomized indicators. Allostatic load scores derived using the distribution-based thresholds were used for the primary analysis.

Our primary analysis used Classification and Regression Trees (CART) [47] to explore allostatic load score thresholds associated with progression-free and overall survival, given an anticipated non-monotonic relationship between allostatic load and these survival outcomes. For each outcome, we fit a univariate tree with allostatic load score alone and a primary multivariate tree including allostatic load, tumor grade, histology, and ethnoracial group. The CART survival analyses yielded hazard ratios, comparing hazard rates of each tree node to the overall analytic sample [48]. Tree complexity was controlled by a complexity parameter (cp=0.005) and a minimum split size of 20 patients. Decision tree analyses including only allostatic load score to model split points for both overall survival and progression-free survival (supplementary figures) were used to select a cut point between “low” and “high” allostatic load scores. Allostatic load was dichotomized ≤3 versus >3 for downstream group comparisons (for “low allostatic load” and “high allostatic load”) based on the convergence of split locations with the decision tree univariable analyses and for a median split approach.

A second set of models were conducted as sensitivity analyses, including separate models using alternative histology classifications, tumor stage, social vulnerability index, and allostatic load scores derived using the clinical thresholds.

All analyses were conducted in R version 4.4.2. Specifically, we used the mice (Multivariate Imputation by Chained Equations) package for multiple imputation, the rpart package for CART decision trees, and the partykit packages for plotting survival curves. All reported *P* values are two-sided, and *P* < 0.05 was considered statistically significant.

## 3. Results

### 3.1 Participant characteristics

As shown in **Table 1**, our study sample comprised majority non-Hispanic White (“White”) women (n = 233), with an equal number of non-Hispanic Black (“Black”, n = 67) and Hispanic (n = 67), and fewer women of other races (n = 31). Hispanic women and women of other races were younger on average (57.1 and 56.4 years, respectively) compared to Black women (62.9 years) and White women (64.2 years). In accordance with oversampling for aggressive disease, 42% of cases were high-grade, with high-grade tumors disproportionately distributed between ethnoracial groups. Over 60% of Black women had high-grade tumors, dissimilar to the proportions among White women (39.9%), Hispanic women (37.3%), and women of other races (25.8%). There were also notable ethnoracial differences in SVI. A majority of Black women (55.2%) and Hispanic women (56.7%) were in the highest SVI tertile, whereas 19.7% of White women and 9.7% of women of other races were in the highest SVI tertile. Median overall and progression-free survival time were not significantly different across ethnoracial groups. Recurrence was reported for 32.8% of Black women, 21.0% of White women, and 19.4% for both Hispanic and other race women.

**Table 1.** Participant characteristics stratified by race/ethnicity group.

|  | Non-Hispanic Black (n=67) | Non-Hispanic White (n=233) | Non-Hispanic Other (n=31) | Hispanic (n=67) | Total (n=398) | p-value |
| --- | --- | --- | --- | --- | --- | --- |
| Age |  |  |  |  |  | < 0.001 |
| Mean (SD) | 62.9 (9.11) | 64.2 (9.85) | 56.4 (12.7) | 57.1 (12.3) | 62.2 (10.8) |  |
| Median [Min, Max] | 64.0 [38.0, 87.0] | 64.0 [29.0, 94.0] | 54.0 [35.0, 84.0] | 58.0 [30.0, 84.0] | 63.0 [29.0, 94.0] |  |
| Missing | 0 (0%) | 0 (0%) | 1 (3.2%) | 0 (0%) | 1 (0.3%) |  |
| Grade |  |  |  |  |  | 0.002 |
| Low grade | 26 (38.8%) | 139 (59.7%) | 23 (74.2%) | 42 (62.7%) | 230 (57.8%) |  |
| High grade | 41 (61.2%) | 93 (39.9%) | 8 (25.8%) | 25 (37.3%) | 167 (42.0%) |  |
| Missing | 0 (0%) | 1 (0.4%) | 0 (0%) | 0 (0%) | 1 (0.3%) |  |
| Histology |  |  |  |  |  | 0.024 |
| Endometrioid | 30 (44.8%) | 153 (65.7%) | 24 (77.4%) | 46 (68.7%) | 253 (63.6%) |  |
| Serous | 17 (25.4%) | 24 (10.3%) | 3 (9.7%) | 8 (11.9%) | 52 (13.1%) |  |
| Carcinosarcoma | 8 (11.9%) | 21 (9.0%) | 1 (3.2%) | 2 (3.0%) | 32 (8.0%) |  |
| Clear cell | 0 (0%) | 6 (2.6%) | 0 (0%) | 1 (1.5%) | 7 (1.8%) |  |
| Mixed | 11 (16.4%) | 26 (11.2%) | 3 (9.7%) | 10 (14.9%) | 50 (12.6%) |  |
| Other (Including missing) | 1 (1.5%) | 3 (1.3%) | 0 (0%) | 0 (0%) | 4 (1.0%) |  |
| Social Vulnerability Index (SVI) |  |  |  |  |  | < 0.001 |
| Mean (SD) | 0.691 (0.261) | 0.446 (0.266) | 0.382 (0.236) | 0.678 (0.305) | 0.521 (0.294) |  |
| Median [Min, Max] | 0.724 [0.0005, 0.997] | 0.440 [0.0003, 0.955] | 0.376 [0.0132, 0.937] | 0.814 [0.0151, 0.997] | 0.529 [0.0003, 0.997] |  |
| Missing | 3 (4.5%) | 14 (6.0%) | 2 (6.5%) | 6 (9.0%) | 25 (6.3%) |  |
| SVI Tertiles |  |  |  |  |  | < 0.001 |
| T1 (<=0.376) | 7 (10.4%) | 92 (39.5%) | 14 (45.2%) | 12 (17.9%) | 125 (31.4%) |  |
| T2 (0.376-0.701) | 20 (29.9%) | 81 (34.8%) | 12 (38.7%) | 11 (16.4%) | 124 (31.2%) |  |
| T3 (>0.701) | 37 (55.2%) | 46 (19.7%) | 3 (9.7%) | 38 (56.7%) | 124 (31.2%) |  |
| Missing | 3 (4.5%) | 14 (6.0%) | 2 (6.5%) | 6 (9.0%) | 25 (6.3%) |  |
| Recurrence |  |  |  |  |  | 0.181 |
| No | 41 (61.2%) | 174 (74.7%) | 22 (71.0%) | 50 (74.6%) | 287 (72.1%) |  |
| Yes | 22 (32.8%) | 49 (21.0%) | 6 (19.4%) | 13 (19.4%) | 90 (22.6%) |  |
| Missing | 4 (6.0%) | 10 (4.3%) | 3 (9.7%) | 4 (6.0%) | 21 (5.3%) |  |
| PFS (Months) |  |  |  |  |  | 0.1 |
| Mean (SD) | 40.2 (23.9) | 40.8 (24.3) | 39.3 (23.9) | 40.5 (25.4) | 40.5 (24.3) |  |
| Median [Min, Max] | 38.8 [5.20, | 38.0 [0.633, | 36.3 [7.73, | 40.9 [0.833, | 38.2 [0.633, |  |
| Missing | 4 (6.0%) | 11 (4.7%) | 3 (9.7%) | 4 (6.0%) | 22 (5.5%) |  |
| OS (Months) |  |  |  |  |  | 0.39 |
| Mean (SD) | 49.0 (24.6) | 43.9 (25.2) | 43.1 (24.3) | 43.3 (25.5) | 44.6 (25.1) |  |
| Median [Min, Max] | 51.4 [1.30, | 45.3 [0.267, | 38.4 [3.00, | 47.7 [0.833, | 47.0 [0.267, |  |
| Missing | 0 (0%) | 0 (0%) | 1 (3.2%) | 0 (0%) | 1 (0.3%) |  |

### 3.2 Allostatic load biomarker heterogeneity and “high risk” thresholds

As shown in **Table 2**, we compared the use of thresholds designated by distribution within our dataset (“distribution-based”) versus standard clinical reference (“clinical”) thresholds to define “high-risk” values in constructing our allostatic load measure. We largely found consistency between distribution-based and standard clinical reference thresholds for high-risk. However, we noted marked differences between distribution-based thresholds and standard clinical thresholds for triglycerides (189 vs. 150 mg/dL), waist circumference (145 vs. 88 cm), CRP (10.3 vs. 3.0 mg/L), IL-6 (8.1 vs. 1.8 pg/mL), and fasting glucose (152.7 vs. 110.0). Notably, based on standard clinical thresholds, values among the women in this sample approached ceiling effects for multiple variables, with >99% exceeding the waist circumference threshold, 91% exceeding the IL-6 threshold, 66% exceeding BMI threshold, 62% exceeding fasting glucose threshold, and 61% exceeding the CRP threshold. As described in the Methods section, we used the distribution-based approach as our primary analysis to maintain the greatest heterogeneity in the allostatic load scoring system.

**Table 2.** Comparison of distribution-based thresholds versus clinical reference range thresholds for “high-risk” on each collected variable.

| Biomarker | Quantile used for distribution | Risk rule for threshold | Distribution threshold | Clinical reference threshold | N at high-risk (distribution-based) | N at high-risk (clinical) |
| --- | --- | --- | --- | --- | --- | --- |
| BP systolic- mmHg | Q3 (75%) | > Q3 | 148.000 | 140.0 | 107 | 174 |
| BP diastolic- mmHg | Q3 (75%) | > Q3 | 81.000 | 90.0 | 108 | 19 |
| HDL- mg/dL | Q1 (25%) | < Q1 | 43.275 | 50.0 | 100 | 169 |
| Total cholesterol mg/dL | Q3 (75%) | > Q3 | 229.495 | 240.0 | 121 | 108 |
| LDL- mg/dL | Q3 (75%) | > Q3 | 128.632 | 130.0 |  |  |
| Triglycerides- mg/dL | Q3 (75%) | > Q3 | 189.027 | 150.0 | 100 | 178 |
| Waist circumference – cm | Q3 (75%) | > Q3 | 145.196 | 88.0 | 100 | 397 |
| BMI – kg/m<sup>2</sup> | Q3 (75%) | > Q3 | 40.663 | 30.0 | 100 | 261 |
| Resting heart rate- bpm | Q3 (75%) | > Q3 | 86.000 | 100.0 | 99 | 28 |
| HbA1c- % | Q3 (75%) | > Q3 | 6.300 | 6.5 | 93 | 79 |
| Albumin- g/dL | Q1 (25%) | < Q1 | 3.950 | 4.0 | 98 | 113 |
| CRP- mg/L | Q3 (75%) | > Q3 | 10.308 | 3.0 | 100 | 241 |
| IL-6- pg/mL | Q3 (75%) | > Q3 | 8.067 | 1.8 | 100 | 362 |
| eGFR | Q1 (25%) | < Q1 | 63.000 | 60.0 | 96 | 84 |
| Creatinine- mg/dL | Q3 (75%) | > Q3 | 1.120 | 1.2 | 98 | 82 |
| Glucose (fasting)- mg/dL | Q3 (75%) | > Q3 | 152.678 | 110.0 | 100 | 245 |

The proportion of women meeting “high-risk” thresholds for SBP, DBP, or HDL levels set by distribution-based cutoffs showed no major ethnoracial differences. However, the proportion of participants with values meeting high-risk threshold levels for triglycerides, BMI, HbA1C, eGFR, and creatinine varied across ethnoracial groups (see supplementary tables).

Cumulative scoring (+1) for each high-risk variable resulted in classifying 43% of the study sample as “low allostatic load” (score ≤3), substantially preserving statistical power (compared to the clinical thresholds approach, which resulted in only 9.7% with allostatic load score ≤3). As shown in **Table 3**, a greater proportion of Black women had high allostatic load scores (68.7%) compared with other groups (56.7%, 56.7%, and 32.3% for White, Hispanic, and other race women, respectively).

**Table 3.** Allostatic load scores (distribution-based) by race/ethnicity groups

|  | Non-Hispanic<br>Black (n=67) | Non-Hispanic<br>White<br>(n=233) | Non-Hispanic<br>Other (n=31) | Hispanic<br>(n=67) | Total<br>(n=398) | p-value |
| --- | --- | --- | --- | --- | --- | --- |
| Allostatic load (AL) |  |  |  |  |  | < 0.001 |
| Mean (SD) | 4.81 (2.02) | 4.11 (2.24) | 2.71 (2.08) | 4.01 (2.19) | 4.10 (2.23) |  |
| Median [Min, Max] | 4.00 [1.00, 9.00] | 4.00 [0, 11.0] | 2.00 [0, 8.00] | 4.00 [0, 11.0] | 4.00 [0, 11.0] |  |
| AL Group |  |  |  |  |  | 0.01 |
| Low (0-3) | 21 (31.3%) | 101 (43.3%) | 21 (67.7%) | 29 (43.3%) | 172 (43.2%) |  |
| High (4-15) | 46 (68.7%) | 132 (56.7%) | 10 (32.3%) | 38 (56.7%) | 226 (56.8%) |  |

### 3.3 Allostatic load and overall survival

We evaluated decision tree models for overall survival, which evaluated covariates for strength of association with survival outcomes. In these analyses, split points for allostatic load score were defined by the model (rather than predetermined in our analysis). In the decision tree models of overall survival, including allostatic load score, tumor grade, endometrioid vs non-endometrioid histology, and race/ethnicity as covariates (**Figure 1**), grade was the primary split, as expected. Among those with low-grade tumors, allostatic load scores of ≥ 8 were associated with a lower survival rate (hazard ratios (HR) = 1.50), compared with HR = 0.54 among those with allostatic load scores ≥2 and <8, and HR = 0.21 among those with the lowest allostatic load scores of <2. In the high-grade tumor group, the highest risk of dying was observed among those with intermediate allostatic load scores (>=4 and <8) and, for those with scores of <4, ethnoracially minoritized participants. A sensitivity analysis including stage at diagnosis was performed and did not substantively change these relationships. Interestingly, an additional sensitivity analysis using 5-category histologic variables (endometrioid, serous, clear cell, carcinosarcoma, and mixed histology) showed that the high grade branching first split by clear cell (HR = 3.7) and then further split by carcinosarcoma versus a combined endometrioid, serous, and mixed histology arm, showing allostatic load score only influencing overall survival for participants that are not Black (sensitivity analysis figures are attached in **supplementary information**).

**Figure 1.**
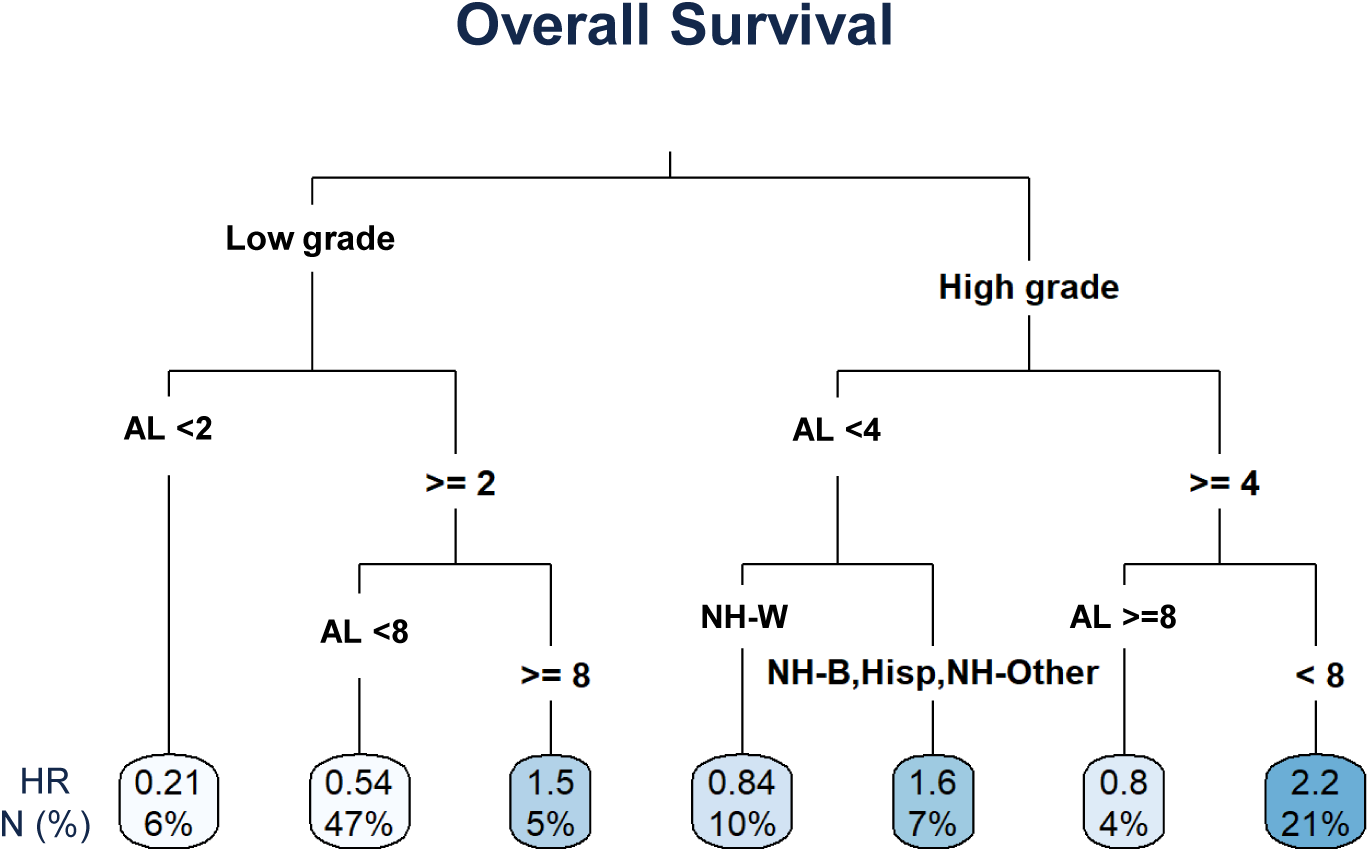
Decision tree analysis for overall survival, including allostatic load (AL) score, tumor grade, histology (endometrioid vs non-endometrioid), and race/ethnicity to indicate hazard ratios (HR) and number of participants (n, %) indicated in each node. HRs are estimated relative to the full sample. Shaded boxes denote increasing HR. (NH-W, non-Hispanic White; NH-B, non-Hispanic Black; NH-Other, non-Hispanic other races.)

### 3.4 Allostatic load and progression free survival

Our second survival tree analysis yielded distinct relationships of allostatic load with progression-free survival based on tumor type and ethnoracial category (**Figure 2**). In the model including allostatic load score, tumor grade, histology (either 2-category or 5-category), and ethnoracial group, the primary split was by grade, separating women into two groups with markedly different patterns of progression-free survival. Among women with low-grade tumors, the ethnoracial group was the only split, with HRs of 1.50, 0.64, and 0.28 among Black women, White women and Hispanic women, and women of other races, respectively. In contrast, among women with high-grade tumors, the first split was on allostatic load, with the rate of disease progression among women with allostatic load score ≥9 being 0.52, which is 48% lower progression rate than the population, with no differences by ethnoracial group. Among those with allostatic load scores <9, distinct survival patterns by ethnoracial group emerged at the allostatic load score ≥4 split. Among those with the lowest allostatic load scores (<4), Black women had twice the rate of disease progression (HR = 2.00). However, for women with allostatic load scores ≥4 and <9, Black women 20% higher progression rate compared to the population (HR = 1.2) but White and other race women had more than more than twice the rate of disease progression (HR = 2.50). Sensitivity analyses, including stage in these models, did not substantively alter the allostatic load score relationships or patterns.

**Figure 2.**
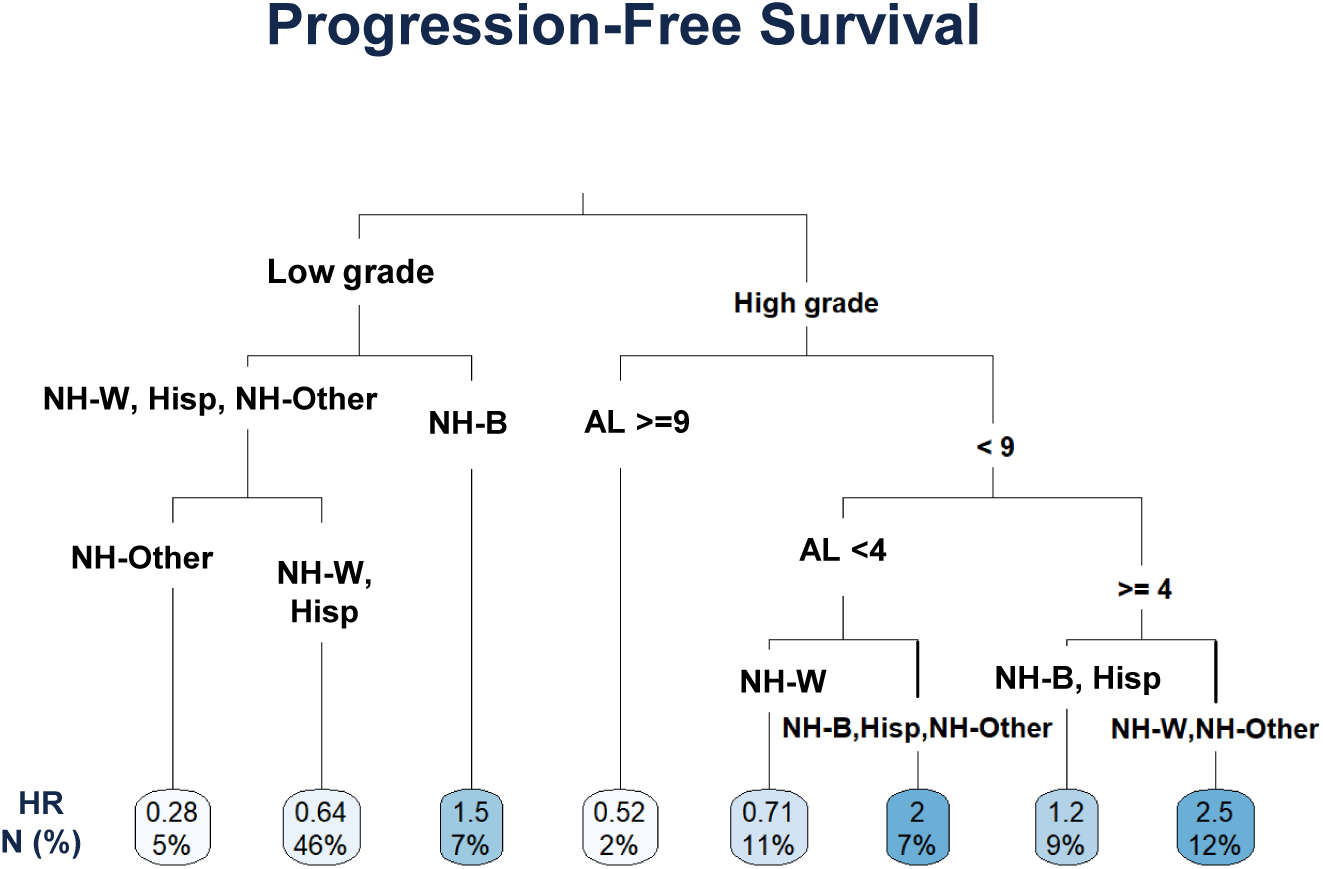
Decision tree analysis for progression-free survival, including allostatic load (AL) score, tumor grade, histology (endometrioid vs non-endometrioid), and race/ethnicity to indicate hazard ratios (HR) and number of participants (n, %) in each node. HRs are estimated relative to the full sample. Shaded boxes denote increasing HR. (NH-W, non-Hispanic White; NH-B, non-Hispanic Black; NH-Other, non-Hispanic other races.)

## 4. Discussion

In this initial exploratory investigation, our results suggest that the relationships of allostatic load with endometrial cancer outcomes vary by tumor grade and ethnoracial group. We observed several notable findings regarding clinical outcomes in this small hypothesis-generating cohort study. First, we found that the association between allostatic load seemed to have the strongest impact on overall survival in low-grade cancers (with increasing HR across increasing allostatic load scores). In contrast, we noted that the relationship between allostatic load score and overall survival was different for women with high-grade endometrial cancers, where survival durations were shortest among those with intermediate allostatic load scores (>=4 and <8) and for ethnoracially minoritized groups with allostatic load scores <4.

Our results suggest distribution-based individual variable thresholds maintain heterogeneity across the measured domains for an obesity-dominant endometrial cancer cohort. Standard clinical thresholds for identifying “high-risk” biomarker values produced substantial ceiling effects, where nearly the entire sample exceeded the standard waist circumference, IL-6, and CRP thresholds, reflecting the predominance of obesity in this population. When using the distribution-based biomarker thresholds, we found allostatic load heterogeneity persists despite the high degree of obesity in this cohort. Although >99% of participants exceeded the clinical waist circumference threshold (or >65% had BMI>30 kg/m^2^), allostatic load score distribution was not overly compressed and retained sufficient variability, confirming that meaningful individual variation in cumulative metabolic and physiologic stress persists even when obesity is near-universal. These results support a central premise in our study: obesity status alone (defined by BMI) does not capture the heterogeneity in cumulative physiologic/metabolic stressors that may underlie disparate endometrial cancer outcomes. This is an important conceptual contribution to the field and further supports our approach to incorporating a cumulative score across physiologic systems (allostatic load) in illuminating physiological mechanisms underpinning endometrial cancer risk.

The relationship between allostatic load and clinical outcomes differed between overall survival and progression-free survival, suggesting a potential interaction between allostatic load and mortality risk beyond cancer recurrence (or progression). Further, we found that the relationship between allostatic load and progression-free survival differed across tumor grade. Among women with low-grade endometrial cancer, progression-free survival was shortest in Black women, irrespective of allostatic load. This finding is consistent with a prior large-sample study identifying higher rates of disease progression among Black compared with White women with low-grade endometrial cancer [49]. The results of our study indicate the persistence of this disparity even when attempting to account for Black women’s higher cumulative physiologic burden. Surprisingly, among women with high-grade endometrial cancer, very high allostatic load (≥9) was associated with *longer* progression-free survival, whereas progression-free survival rate depended on race and ethnicity among the women with the lowest allostatic load scores.

Elevated allostatic load may adversely influence progression-free and overall survival in endometrial cancer through multiple interconnected biological and behavioral pathways. Chronic physiological stress is associated with dysregulation of the neuroendocrine, metabolic, cardiovascular, and immune systems, resulting in sustained inflammation, impaired immune surveillance, and altered hormonal signaling that may promote tumor growth and metastatic progression [9, 50–54]. Persistent activation of stress-response pathways, including the hypothalamic–pituitary–adrenal axis and sympathetic nervous system, can increase circulating inflammatory mediators and glucocorticoids, contributing to a tumor microenvironment that facilitates angiogenesis, cellular proliferation, and resistance to apoptosis [1, 55–57]. Components of allostatic load such as obesity, insulin resistance, hypertension, and diabetes are also independently associated with poorer endometrial cancer outcomes and may further exacerbate disease progression through metabolic and inflammatory mechanisms [58–62]. In addition to these direct biological effects, high allostatic load may reflect cumulative social and structural disadvantage that influences cancer survival indirectly through delayed diagnosis, decreased access to high-quality care, lower rates of treatment compliance, and greater comorbidity burden, all of which may compromise treatment effectiveness and increase mortality risk [63–67].

Our results indicate that the relationship of allostatic load with endometrial cancer outcomes may be particularly influenced by social processes (upstream or downstream), even for patients that were engaged with treatment at a tertiary care center. One potential contributor for our observation of unexpected impact of very high allostatic load scores on survival is that women with elevated allostatic load may have higher overall morbidity, leading to more frequent engagement with the healthcare system and potentially increased interventions or greater access and adherence to treatment for endometrial cancer, irrespective of race or ethnicity. Conversely, Black and Hispanic women in this cohort with low allostatic load but high-grade tumors may have fewer routine healthcare interactions. In these groups, structural barriers, experiences of discrimination, and higher levels of healthcare mistrust may further limit timely access to care [68–71] or receipt of adjuvant treatment [72–74], which is especially critical for high-grade disease [66], potentially contributing to shorter progression-free survival at low levels of allostatic load in these groups.

In contrast, White women in this cohort with high-grade endometrial cancer but generally low allostatic load may be in better overall health. If low allostatic load is a surrogate indicator for overall higher SES, these individuals may experience fewer barriers to care, which may explain their relatively better outcomes among the low allostatic load group. We also observed that Black and Hispanic women with mid-range allostatic load scores (≥4, <9) experienced longer progression-free survival than White women with comparable scores. At certain thresholds of morbidity, ethnoracially marginalized women may be less deterred from seeking healthcare for their cumulative burden of chronic conditions than their counterparts with better overall health, thereby yielding more frequent contact with healthcare systems, and increasing opportunities for disease monitoring, management of comorbidities, and continuity of oncologic care that could contribute to prolonged progression-free survival [66].

Notably, our findings also support that intermediate allostatic load scores (i.e., allostatic load scores ≥4, <9) may identify a distinct high-risk subgroup among White women, for whom we observed the shortest progression-free survival. In this group, the physiological dysregulation reflected by elevated allostatic load may contribute directly to disease progression through cardiovascular, metabolic, immune, and inflammatory pathways. At the same time, White women with higher morbidity may also be disproportionately affected by structural barriers to timely and high-quality care relative to their healthier counterparts [75–77]. Prior studies suggest that White women with greater comorbidity burden are more likely to have lower socioeconomic status and reside in rural communities [75], factors also associated with reduced healthcare access and continuity of care. Thus, moderate elevations in allostatic load in White women may represent a clinical profile characterized by sufficient physiological burden to adversely affect cancer outcomes, but without the level of illness severity that reliably prompts sustained engagement with healthcare services. Because Black women are substantially more likely to be diagnosed with high-grade endometrial cancer, these findings are consistent with the broader literature identifying healthcare access and quality as major drivers of ethnoracial disparities in endometrial cancer outcomes.

Our findings partially align with prior work linking allostatic load to tumor clinicopathologic characteristics and survival in ovarian [78] and breast [79] cancers. To our knowledge, this is the first manuscript to examine the relationship between allostatic load and endometrial cancer, and only the second empirical study to evaluate allostatic load in relation to tumor characteristics and outcomes in a specific cancer type. Interestingly, no other study has shown longer survival outcomes associated with higher allostatic load in specific patient subgroups or by tumor type. (In breast cancer, higher allostatic load has been associated with poorer prognostic characteristics.) The variable impact of higher allostatic load across tumor types and ethnoracial groups may reflect the intersecting nature of multiple obesity-associated medical comorbidities and obesity-related healthcare barriers, and warrants further study. These distinctions between our findings and those linking higher allostatic load to poorer breast and ovarian cancer outcomes may reflect, in part, the high prevalence of obesity and/or other metabolic dysregulations among women with endometrial cancer.

This study has several important strengths. To our knowledge, it is the first to characterize the performance of allostatic load scoring in this obesity-centered cancer context and to examine the association between allostatic load and survival outcomes in endometrial cancer. This study extends the literature on physiological stress and cancer disparities into a malignancy characterized by substantial ethnoracial inequities in diagnosis and outcomes. Despite limited sample size, the inclusion of Black, Hispanic, and White women in this study enabled assessment of associations across groups that are frequently underrepresented in oncologic research and provided an opportunity to explore potential ethnoracial differences in the relationship between allostatic load and survival outcomes. In addition, the use of a multidimensional allostatic load measure allowed for the evaluation of cumulative physiological dysregulation across multiple biological systems rather than reliance on any single comorbidity or biomarker.

Several limitations should also be considered. The relatively small sample size limited statistical power, particularly for formal analyses of effect measure modification by race and ethnicity, and subgroup estimates should therefore be interpreted cautiously. Limited sample size may also have reduced the precision of survival estimates within stratified analyses, and precluded analyses among women of other ethnoracial backgrounds beyond Black, White, and Hispanic women. Confidence intervals were not reported for HRs derived from CART survival analyses because terminal-node estimates are based on small, data-dependent subsamples, and the tree structure itself may vary across samples. It cannot be excluded that progression-free survival could be falsely extended for patients that experience barriers to timely follow-up to detect disease progression/recurrence, and these barriers could be impacted by differing degrees of medical comorbidities in these groups. In addition, observations within terminal nodes are not independent and identically distributed in the usual inferential sense, making standard confidence interval estimation for node-specific hazard ratios inappropriate [48].

Because allostatic load indices are not standardized across studies, the specific biomarker composition and thresholds used in this analysis may affect comparability with other populations and settings. Our operationalization of allostatic load was limited to the biomarkers collected within the cohort, which did not include neuroendocrine markers. Thus, while our allostatic load measure represented multiple physiological systems, our analysis did not consider the role that neuroendocrine mechanisms may play in driving endometrial cancer outcomes. As this study analyzed allostatic load in endometrial cancer patients seeking care at an academic tertiary care center (MD Anderson Cancer Center), participants may not represent local and regional patient characteristics; however, SVI scores reflected moderately-high social vulnerability in this cohort. This patient cohort sought care in Houston, Texas, which will also reflect spatial, social, and cultural factors unique to this area and is not nationally representative. Participants in this study had been previously consented for banking blood and tissue specimens for future research purposes, which could bias the population by excluding those who were unable to undergo surgery or based on willingness to participate in research. Finally, as with all observational studies, residual confounding and potential selection biases cannot be excluded.

The findings of this study highlight potential pathways contributing to persistent racial and ethnic disparities in survival among women with endometrial cancer and underscore the importance of considering both cumulative physiological stress and structural determinants of health inequities in studies of cancer outcomes. Although higher allostatic load was generally associated with poorer progression-free and overall survival, our exploratory analyses suggest that the relationship between physiological burden and survival is not uniform across racial and ethnic groups or tumor subtypes. Rather, the observed heterogeneity indicates that allostatic load may operate within broader social and clinical contexts that shape access to care, healthcare utilization, treatment continuity, and comorbidity management. These findings support the need for multidimensional frameworks that integrate biological, behavioral, and structural pathways when examining disparities in endometrial cancer outcomes.

Importantly, the observation that Black women with low allostatic load and low-grade disease nevertheless experienced poorer progression-free survival suggests that reducing physiological stress burden alone may be insufficient to eliminate disparities in cancer outcomes. Such findings are consistent with a growing literature demonstrating that differential exposure to chronic stressors and differential effects of those stressors may coexist alongside preventable differences in healthcare access, quality of care, treatment delays, symptom recognition, and clinician response [66, 80]. In this context, allostatic load may function not only as a biological marker of cumulative stress exposure, but also as an indicator of broader social and structural factors that interacts with tumor biology and healthcare systems to shape prognosis.

Our findings also raise important questions regarding the clinical interpretation of allostatic load in endometrial cancer populations. The non-linear associations observed in the survival tree analyses, particularly among women with high-grade tumors, suggest that specific allostatic load thresholds may identify clinically distinct subgroups with differing patterns of disease vulnerability and possibly differences in healthcare engagement, which should be further defined in future studies. Future studies should investigate whether moderate versus severe elevations in allostatic load correspond to differences in healthcare utilization, treatment adherence, comorbidity management, or surveillance intensity. Longitudinal analyses incorporating treatment data, healthcare access measures, and trajectories of physiologic dysregulation over time may help clarify the mechanisms underlying these relationships. Both systemic and endometrial tissue-level resilience factors (antioxidant and cytoprotective responses) are also critical factors that should be investigated in future studies of both cancer risk and tumor metabolic programs.

Further research is also needed to evaluate the biological pathways linking allostatic load with endometrial cancer progression. Given the established roles of inflammation, obesity, metabolic dysfunction, and endocrine dysregulation in endometrial carcinogenesis, future studies integrating molecular tumor profiling, therapeutic regimen details, inflammatory biomarkers, and measures of stress physiology may help identify mechanistic pathways through which chronic stress influences tumor progression and survival. Such work may be particularly important for understanding the intersection of obesity-related metabolic dysfunction and cumulative physiological stress in endometrial cancer, a disease strongly associated with cardiometabolic comorbidity.

Finally, replication in larger and more geographically diverse cohorts is essential. Larger studies would permit formal testing of effect measure modification by ethnoracial group, tumor characteristics, socioeconomic status, and healthcare access variables, while also enabling validation of clinically meaningful allostatic load thresholds. Allostatic load may represent a valuable multidimensional marker for identifying patients at elevated risk of adverse outcomes who could benefit from enhanced supportive care, chronic disease management, and interventions addressing barriers to timely and equitable cancer care.

## Statements and Declarations

### Funding

This work was supported in part by National Cancer Institute (NCI) U54 CA302426 and MD Anderson Institutional Research Grant Seed Funding to MSY, Cancer Prevention & Research Institute of Texas Proteomics & Metabolomics Core Facility Support Award (RP210227), and NCI Cancer Center Support Grant (P30CA125123) to the Antibody-based Proteomics Core/Shared Resource at Dan L. Duncan Cancer Center at Baylor College of Medicine.

### Competing Interests

GSB, MBB, SW, MB, LB, QZ, BM, JC, CS, FCL, and MSY have no relevant financial or non-financial interests to disclose. Outside the scope of this study, JAB serves as Consultant for Abbott Nutrition and holds equity in SynchroHealth LLC (Chief Scientific Officer).

### Author Contributions

MBB, LB, and MSY contributed to the study conceptualization and design. GSB, SW, MB, CS, FCL contributed to formal analysis, methodology, and data visualization. JAB contributed to interpretation of results and manuscript review. QZ, BM, LB, MBB, and JC contributed to material preparation and data collection. GSB, MBB, and MSY contributed first drafts of the manuscript. All authors reviewed and commented on the earlier drafts and approve of the final submitted version. In addition to points above, MSY led study supervision, project administration, and funding acquisition.

### Ethics Approvals

This study was performed in line with the principles of the Declaration of Helsinki. Approval was granted by the Institutional Review Board of MD Anderson Cancer Center and University of North Carolina at Chapel Hill. All participants provided informed consent for inclusion of biospecimens and data in the MD Anderson Gynecologic Oncology Tumor Bank.

### Data availability

The data that support the findings of this study are not openly available to protect study participant privacy but limited deidentified information may be available through request from the corresponding author upon reasonable request and under permission from MD Anderson Cancer Center.

## Acknowledgments

This work was supported in part by National Cancer Institute (NCI) U54 CA302426 and MD Anderson Institutional Research Grant Seed Funding to MSY, Cancer Prevention & Research Institute of Texas Proteomics & Metabolomics Core Facility Support Award (RP210227), and NCI Cancer Center Support Grant (P30CA125123) to the Antibody-based Proteomics Core/Shared Resource at Dan L. Duncan Cancer Center at Baylor College of Medicine. M.B.B. was supported by a predoctoral fellowship from the Cancer Prevention and Research Institute of Texas (RP210042). We wish to acknowledge Shixia Huang, Zhongcheng Shi, and Michael Nguyen for their excellent technical assistance in performing Luminex experiments, data preliminary analyses and QC, and project consultation. Manuscript content is solely the responsibility of the authors and does not necessarily represent the official views of the National Institutes of Health or other funding agencies.

We gratefully acknowledge the late Diana Urbauer, Principal Biostatistician at MD Anderson, for her important contributions to the statistical analysis plan and IRB protocol development for this study. Diana was a cherished colleague with an unwavering commitment to upholding the most rigorous expectations for study design and analyses on behalf of our current and future patients.

## Notes

### Author Declarations

Institutional Review Boards at the University of Texas MD Anderson Cancer Center gave ethical approval for this work. Institutional Review Boards at the University of North Carolina School of Medicine gave ethical approval for this work.

