## Supplementary for "Allostatic Load in Endometrial Cancer Disparities"

Supplementary Table 1. Comparison of “high-risk” values by individual biomarker across race/ethnicity groups using distribution-based thresholds

| | Non-Hispanic Black (n=67) | Non-Hispanic White (n=233) | Non-Hispanic Other (n=31) | Hispanic (n=67) | Total (n=398) | p-value |
|---|---|---|---|---|---|---|
| SBP > 148 mmHg | | | | | | 0.797 |
| No | 49 (73.1%) | 168 (72.1%) | 25 (80.6%) | 49 (73.1%) | 291 (73.1%) | |
| Yes | 18 (26.9%) | 65 (27.9%) | 6 (19.4%) | 18 (26.9%) | 107 (26.9%) | |
| DBP > 81 mmHg | | | | | | 0.896 |
| No | 48 (71.6%) | 171 (73.4%) | 21 (67.7%) | 50 (74.6%) | 290 (72.9%) | |
| Yes | 19 (28.4%) | 62 (26.6%) | 10 (32.3%) | 17 (25.4%) | 108 (27.1%) | |
| HDL < 43.3 mg/dL | | | | | | 0.164 |
| No | 51 (76.1%) | 181 (77.7%) | 23 (74.2%) | 43 (64.2%) | 298 (74.9%) | |
| Yes | 16 (23.9%) | 52 (22.3%) | 8 (25.8%) | 24 (35.8%) | 100 (25.1%) | |
| Total Chol and LPL | | | | | | 0.162 |
| No | 50 (74.6%) | 152 (65.2%) | 24 (77.4%) | 51 (76.1%) | 277 (69.6%) | |
| Yes | 17 (25.4%) | 81 (34.8%) | 7 (22.6%) | 16 (23.9%) | 121 (30.4%) | |
| Triglycerides > 189 | | | | | | 0.001 |
| No | 60 (89.6%) | 171 (73.4%) | 26 (83.9%) | 41 (61.2%) | 298 (74.9%) | |
| Yes | 7 (10.4%) | 62 (26.6%) | 5 (16.1%) | 26 (38.8%) | 100 (25.1%) | |
| Waist (girth) > 145 cm | | | | | | 0.501 |
| No | 48 (71.6%) | 171 (73.4%) | 25 (80.6%) | 54 (80.6%) | 298 (74.9%) | |
| Yes | 19 (28.4%) | 62 (26.6%) | 6 (19.4%) | 13 (19.4%) | 100 (25.1%) | |
| BMI > 40.7 kg/m$^2$ | | | | | | 0.002 |
| No | 40 (59.7%) | 177 (76.0%) | 29 (93.5%) | 52 (77.6%) | 298 (74.9%) | |
| Yes | 27 (40.3%) | 56 (24.0%) | 2 (6.5%) | 15 (22.4%) | 100 (25.1%) | |
| Resting heart rate > 86 | | | | | | 0.047 |
| No | 49 (73.1%) | 166 (71.2%) | 27 (87.1%) | 57 (85.1%) | 299 (75.1%) | |
| Yes | 18 (26.9%) | 67 (28.8%) | 4 (12.9%) | 10 (14.9%) | 99 (24.9%) | |
| HbA1C > 6.3 % | | | | | | 0.007 |
| No | 46 (68.7%) | 190 (81.5%) | 26 (83.9%) | 43 (64.2%) | 305 (76.6%) | |
| Yes | 21 (31.3%) | 43 (18.5%) | 5 (16.1%) | 24 (35.8%) | 93 (23.4%) | |
| Albumin < 3.95 g/dL | | | | | | 0.879 |
| No | 48 (71.6%) | 178 (76.4%) | 23 (74.2%) | 51 (76.1%) | 300 (75.4%) | |
| Yes | 19 (28.4%) | 55 (23.6%) | 8 (25.8%) | 16 (23.9%) | 98 (24.6%) | |
| CRP > 10.3 mg/L | | | | | | 0.075 |
| No | 47 (70.1%) | 174 (74.7%) | 29 (93.5%) | 48 (71.6%) | 298 (74.9%) | |
| Yes | 20 (29.9%) | 59 (25.3%) | 2 (6.5%) | 19 (28.4%) | 100 (25.1%) | |
| IL6 > 8.07 pg/mL | | | | | | 0.097 |

| | | | | | | |
|---|---|---|---|---|---|---|
| No | 49 (73.1%) | 170 (73.0%) | 29 (93.5%) | 50 (74.6%) | 298 (74.9%) | |
| Yes | 18 (26.9%) | 63 (27.0%) | 2 (6.5%) | 17 (25.4%) | 100 (25.1%) | |
| eGFR < 63 | | | | | | < 0.001 |
| No | 36 (53.7%) | 180 (77.3%) | 29 (93.5%) | 57 (85.1%) | 302 (75.9%) | |
| Yes | 31 (46.3%) | 53 (22.7%) | 2 (6.5%) | 10 (14.9%) | 96 (24.1%) | |
| Creatinine > 1.12 mg/dL | | | | | | < 0.001 |
| No | 35 (52.2%) | 180 (77.3%) | 28 (90.3%) | 57 (85.1%) | 300 (75.4%) | |
| Yes | 32 (47.8%) | 53 (22.7%) | 3 (9.7%) | 10 (14.9%) | 98 (24.6%) | |
| Fasting glucose > 152.7 | | | | | | 0.47 |
| No | 53 (79.1%) | 170 (73.0%) | 26 (83.9%) | 49 (73.1%) | 298 (74.9%) | |
| Yes | 14 (20.9%) | 63 (27.0%) | 5 (16.1%) | 18 (26.9%) | 100 (25.1%) | |
| Medication indicator | | | | | | 0.185 |
| No | 41 (61.2%) | 172 (73.8%) | 22 (71.0%) | 51 (76.1%) | 286 (71.9%) | |
| Yes | 26 (38.8%) | 61 (26.2%) | 9 (29.0%) | 16 (23.9%) | 112 (28.1%) | |

Supplementary Table 2. Descriptive statistics by allostatic load group

| | Low AL (0-3) (N=172) | High AL (4-15) (N=226) | Total (N=398) | p-value |
|---|---|---|---|---|
| Age, mean (SD) | 60.7 (10.6) | 63.3 (10.9) | 62.2 (10.8) | 0.016 |
| Median [Min, Max] | 61.0 [30.0, 88.0] | 64.0 [29.0, 94.0] | 63.0 [29.0, 94.0] | |
| Missing | 1 (0.6%) | 0 (0%) | 1 (0.3%) | |
| Race/ethnicity, % | | | | 0.01 |
| Non-Hispanic Black | 21 (12.2%) | 46 (20.4%) | 67 (16.8%) | |
| Non-Hispanic White | 101 (58.7%) | 132 (58.4%) | 233 (58.5%) | |
| Non-Hispanic Other | 21 (12.2%) | 10 (4.4%) | 31 (7.8%) | |
| Hispanic | 29 (16.9%) | 38 (16.8%) | 67 (16.8%) | |
| SVI, mean (SD) | 0.462 (0.291) | 0.566 (0.288) | 0.521 (0.294) | 0.001 |
| Median [Min, Max] | 0.452 [0.000300, 0.983] | 0.601 [0.000500, 0.997] | 0.529 [0.000300, 0.997] | |
| Missing | 11 (6.4%) | 14 (6.2%) | 25 (6.3%) | |
| Stage | | | | 0.766 |
| Stage I | 121 (70.3%) | 159 (70.4%) | 280 (70.4%) | |
| Stage II | 12 (7.0%) | 12 (5.3%) | 24 (6.0%) | |
| Stage III | 30 (17.4%) | 42 (18.6%) | 72 (18.1%) | |
| Missing | 9 (5.2%) | 13 (5.8%) | 22 (5.5%) | |
| Grade | | | | 0.558 |
| Low grade | 103 (59.9%) | 127 (56.2%) | 230 (57.8%) | |
| High grade | 69 (40.1%) | 98 (43.4%) | 167 (42.0%) | |
| Missing | 0 (0%) | 1 (0.4%) | 1 (0.3%) | |
| Histology | | | | 0.005 |
| Endometrioid | 114 (66.3%) | 139 (61.5%) | 253 (63.6%) | |
| Serous | 30 (17.4%) | 22 (9.7%) | 52 (13.1%) | |
| Other | 28 (16.3%) | 63 (27.9%) | 91 (22.9%) | |
| Missing | 0 (0%) | 2 (0.9%) | 2 (0.5%) | |
| Recurrence | | | | 1 |
| No | 126 (73.3%) | 161 (71.2%) | 287 (72.1%) | |
| Yes | 39 (22.7%) | 51 (22.6%) | 90 (22.6%) | |
| Missing | 7 (4.1%) | 14 (6.2%) | 21 (5.3%) | |
| PFS (Months) | | | | 0.2 |
| Mean (SD) | 42.5 (24.0) | 39.0 (24.5) | 40.5 (24.3) | |
| Median [Min, Max] | 37.9 [0.633, 96.1] | 39.1 [0.733, 98.3] | 38.2 [0.633, 98.3] | |
| Missing | 8 (4.7%) | 14 (6.2%) | 22 (5.5%) | |

| | | | | |
|---|---|---|---|---|
| OS (Months) | | | | 0.18 |
| Mean (SD) | 46.2 (23.9) | 43.4 (25.9) | 44.6 (25.1) | |
| Median [Min, Max] | 48.5 [0.633, 96.1] | 45.8 [0.267, 102] | 47.0 [0.267, 102] | |
| Missing | 1 (0.6%) | 0 (0%) | 1 (0.3%) | |

AL, allostatic load; OS, overall survival; PFS, progression-free survival; SD, standard deviation

### Supplementary Figures

#### OS Tree including AL, grade, race/ethnicity, histology (5-category)

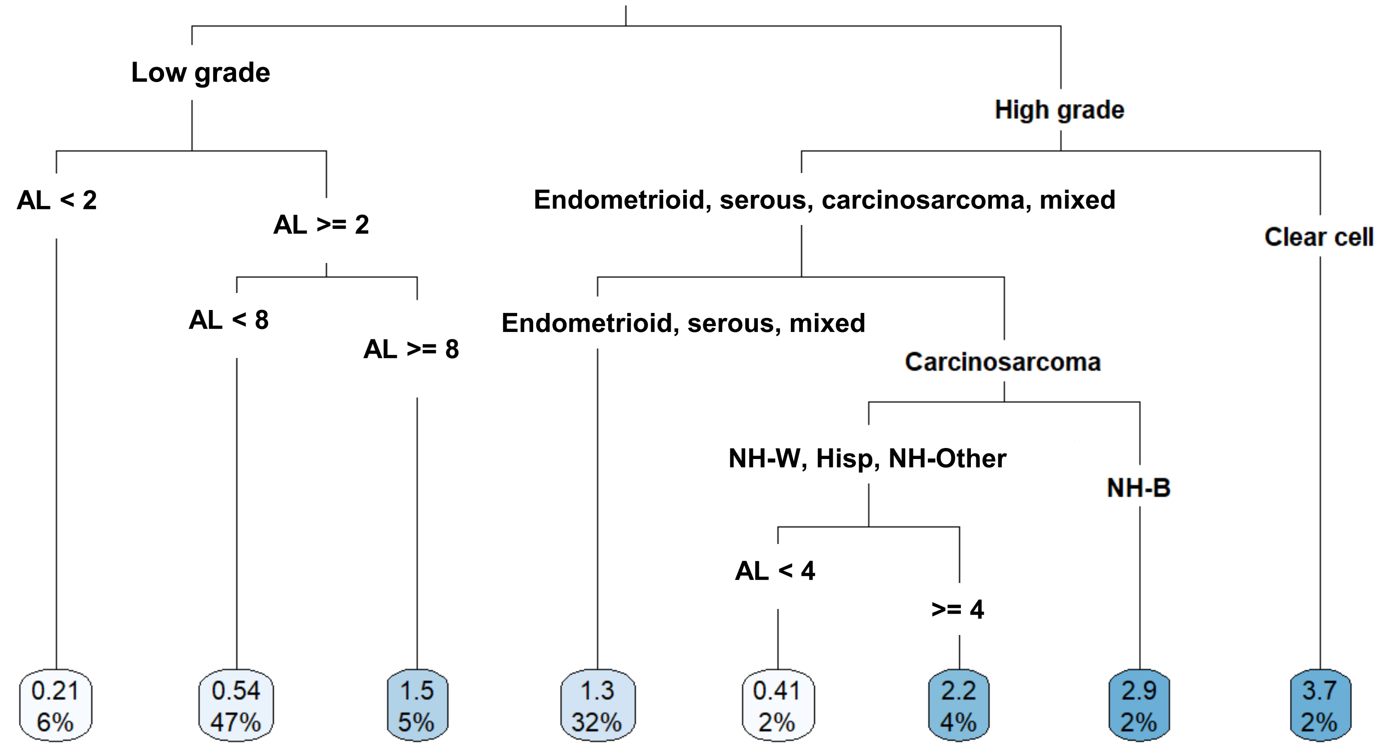

#### Supplementary Figures

### OS Tree including AL, grade, stage, race/ethnicity, histology (3-category)

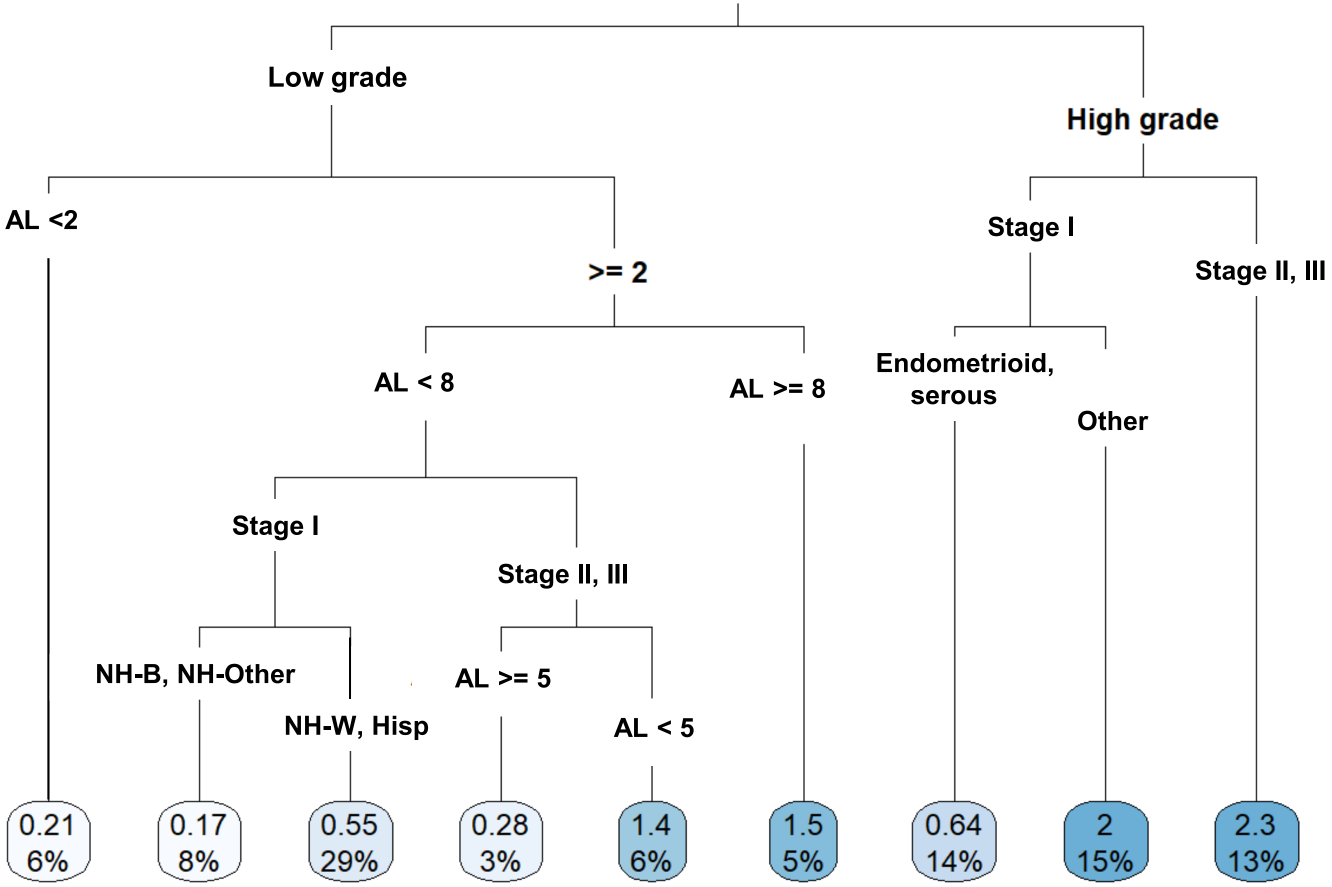

### Supplementary Figures

#### PFS Tree including AL, grade, race/ethnicity, histology (5-category)

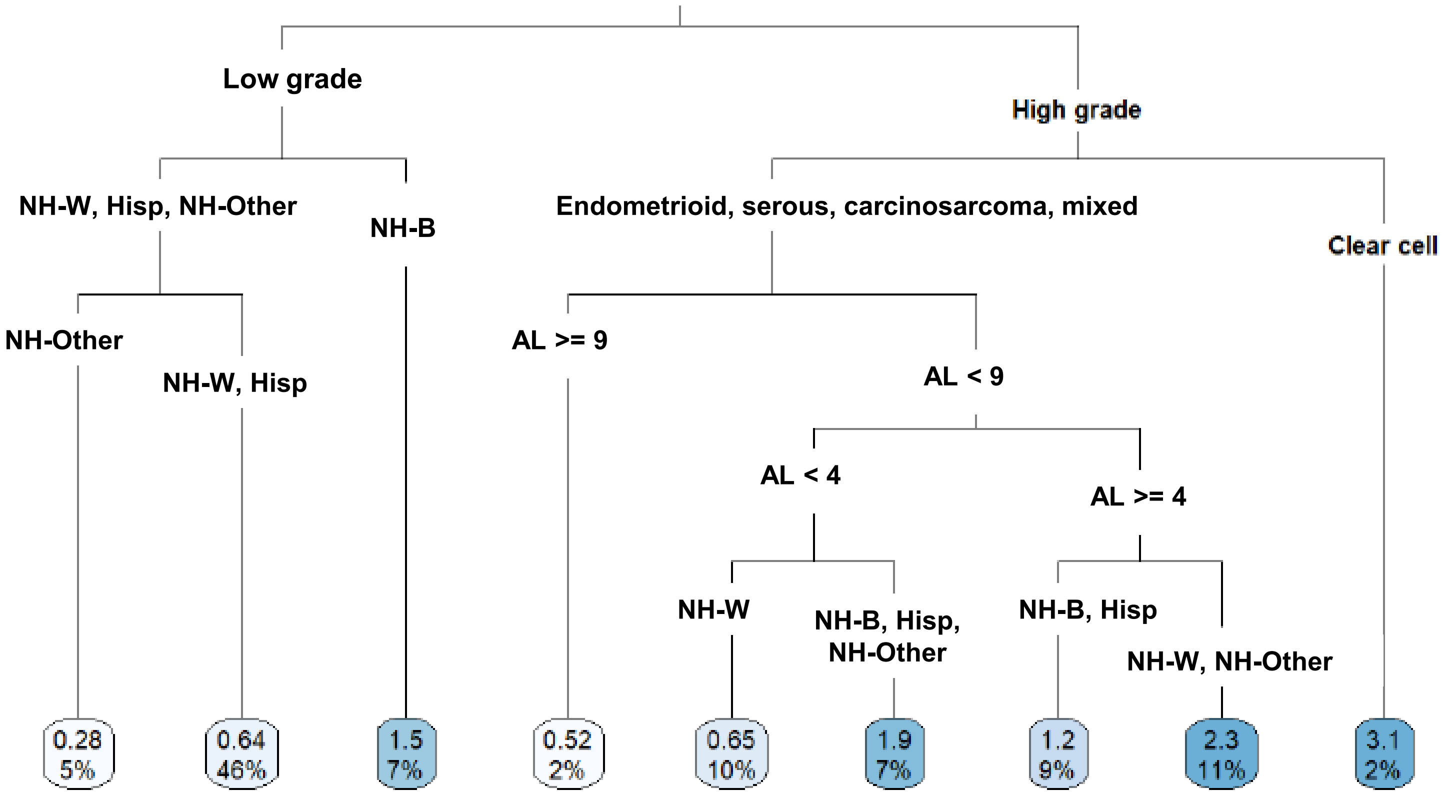

### Supplementary Figures

#### PFS Tree including AL, grade, stage, race/ethnicity, histology (3-category)

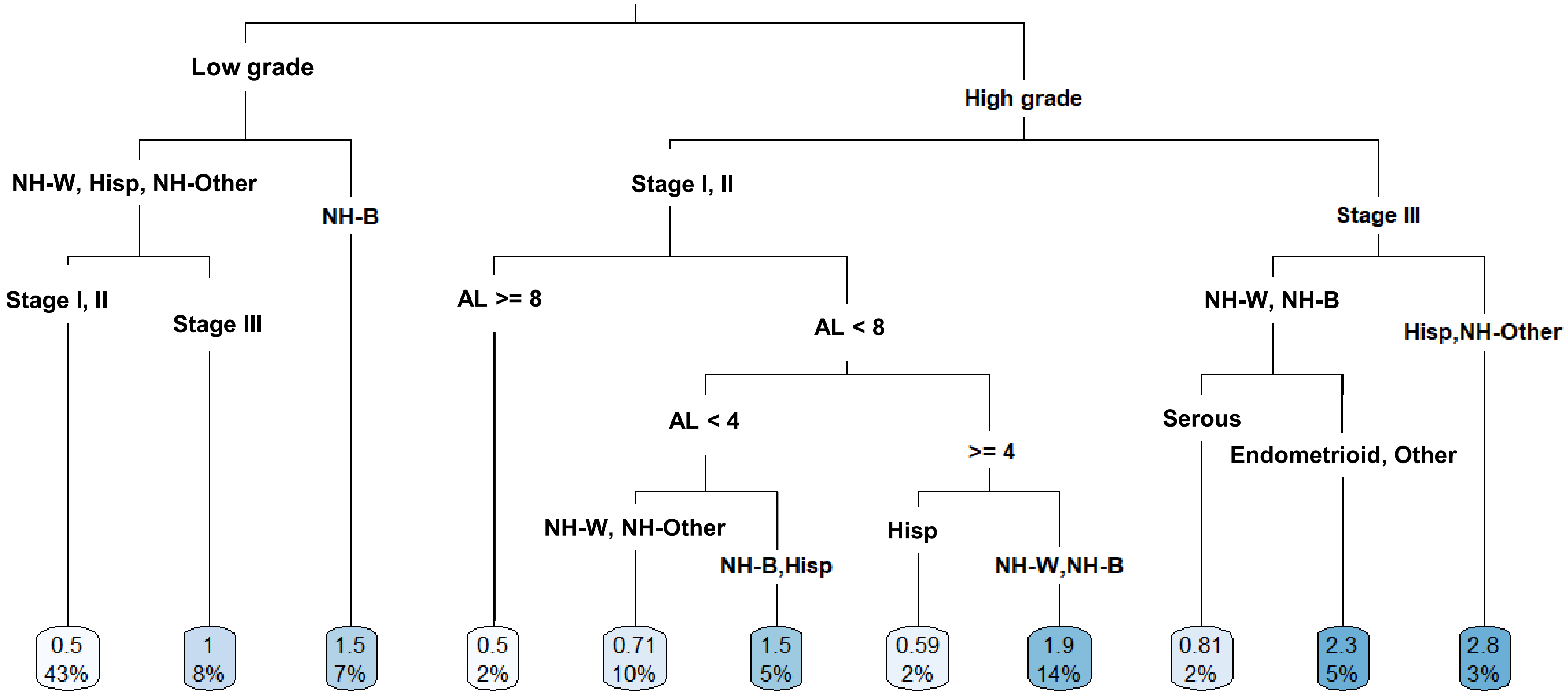
